# Characteristics and Outcomes of COVID-19 Patients in New York City’s Public Hospital System

**DOI:** 10.1101/2020.05.29.20086645

**Authors:** Roopa Kalyanaraman Marcello, Johanna Dolle, Sheila Grami, Richard Adule, Zeyu Li, Kathleen Tatem, Chinyere Anyaogu, Stephen Apfelroth, Raji Ayinla, Noella Boma, Terence Brady, Braulio F. Cosme-Thormann, Roseann Costarella, Kenra Ford, Kecia Gaither, Jessica Jacobson, Marc Kanter, Stuart Kessler, Ross B. Kristal, Joseph J. Lieber, Vikramjit Mukherjee, Vincent Rizzo, Madden Rowell, David Stevens, Elana Sydney, Andrew Wallach, Dave A. Chokshi, Nichola Davis, New York City Health + Hospitals COVID-19 Population Health Data Team

## Abstract

**Background:** New York City (NYC) has borne the greatest burden of COVID-19 in the United States, but information about characteristics and outcomes of racially/ethnically diverse individuals tested and hospitalized for COVID-19 remains limited. In this case series, we describe characteristics and outcomes of patients tested for and hospitalized with COVID-19 in New York City’s public hospital system.

**Methods:** We reviewed the electronic health records of all patients who received a SARS-CoV-2 test between March 5 and April 9, 2020, with follow up through April 16, 2020. The primary outcomes were a positive test, hospitalization, and death. Demographics and comorbidities were also assessed.

**Results:** 22254 patients were tested for SARS-CoV-2. 13442 (61%) were positive; among those, the median age was 52.7 years (interquartile range [IQR] 39.5-64.5), 7481 (56%) were male, 3518 (26%) were Black, and 4593 (34%) were Hispanic. Nearly half (4669, 46%) had at least one chronic disease (27% diabetes, 30% hypertension, and 21% cardiovascular disease). Of those testing positive, 6248 (46%) were hospitalized. The median age was 61.6 years (IQR 49.7-72.9); 3851 (62%) were male, 1950 (31%) were Black, and 2102 (34%) were Hispanic. More than half (3269, 53%) had at least one chronic disease (33% diabetes, 37% hypertension, 24% cardiovascular disease, 11% chronic kidney disease). 1724 (28%) hospitalized patients died. The median age was 71.0 years (IQR 60.0, 80.9); 1087 (63%) were male, 506 (29%) were Black, and 528 (31%) were Hispanic. Chronic diseases were common (35% diabetes, 37% hypertension, 28% cardiovascular disease, 15% chronic kidney disease). Male sex, older age, diabetes, cardiac history, and chronic kidney disease were significantly associated with testing positive, hospitalization, and death. Racial/ethnic disparities were observed across all outcomes.

**Conclusions and Relevance:** This is the largest and most racially/ethnically diverse case series of patients tested and hospitalized for COVID-19 in the United States to date. Our findings highlight disparities in outcomes that can inform prevention and testing recommendations.

## Introduction

SARS-CoV-2 (COVID-19) emerged in December 2019 in Wuhan, China, and it rapidly spread across the world. The United States currently has the most COVID-19 cases globally, and New York City (NYC) is the most affected region, with more than 184000 confirmed cases and more than 15100 confirmed deaths as of May 12, 2020.^1^ The city’s public hospital system has cared for a disproportionate number of cases, with many of its hospitals having neared capacity and having needed additional support.

Reports published to date on COVID-19 patients in NYC have been of cohorts within private health systems that typically serve primarily insured, higher-income patients.^2,3,4,5,6,7^ Additionally, these reports primarily present data only on individuals who have tested positive or have been hospitalized; analyses of characteristics of all patients presenting for testing and their sequelae are lacking. Furthermore, data are limited on patients reflecting the full racial and ethnic diversity of NYC and a wider spectrum of socioeconomic statuses. Identifying characteristics that are associated with a positive test result, hospitalization, and death can help refine public health policy and improve care for groups who are at highest risk of illness and hospitalization.

We describe here the results of our analysis of the demographic and clinical characteristics and outcomes of patients tested for COVID-19 and admitted to New York City’s public hospital system, the largest case series in NYC and the United States reported to date.

## Methods

### Study Setting

New York City Health + Hospitals (NYC H+H) is the largest public health care system in the country. It provides inpatient, outpatient, and home- and community-based services to more than 1 million New Yorkers each year at more than 70 facilities across the city’s five boroughs. The majority of NYC H+H’s patients are low-income: one third (32%) are uninsured and another one third (35%) are Medicaid beneficiaries. Additionally, more than 70% of patients are people of color, many of whom are immigrants. The system includes 11 acute care hospitals, which were the primary setting for this study.

### Data Sources

We extracted medical records for all patients tested for SARS-CoV-2 at any NYC H+H location between March 5 and April 9, 2020. Patients were followed up for primary outcomes (test result, hospitalization, and in-hospital death) through April 16, 2020.

Test results were based on real-time reverse transcriptase polymerase chain reaction (RT-PCR) assays of nasopharyngeal swabs. Initially, assays were conducted by the New York City Department of Health and Mental Hygiene and LabCorp. Starting on March 19, 2020, all tests were processed by BioReference Laboratories. Starting on April 1, 2020, NYC H+H performed some tests using the Cepheid GeneXpert Express SARS-CoV-2 assay. COVID-19 patients were defined as those with a laboratory-confirmed positive test. Hospitalized patients were defined as those who were admitted on or after the date of their test. Patients who died were defined as those with a death recorded in the inpatient setting.

Data on age, sex, race/ethnicity, BMI (calculated from most recent height and weight), and comorbidities were extracted from patients’ electronic health record (EHR) where available. Comorbidities were defined as the presence or absence of the following chronic conditions recorded as billing diagnoses or “active” in a patient’s problem list in the EHR: diabetes, hypertension (HTN), arrhythmia, cardiovascular disease (CVD), congestive heart failure (CHF), asthma, chronic obstructive pulmonary disease (COPD), chronic kidney disease (CKD), liver disease, cancer, HIV, and a flag for having one or more specified chronic disease (diabetes, HTN, CVD, asthma, COPD, or CKD). We selected these based on prior published reports and current clinical experience with COVID-19 patients at NYC H+H.^2,8,9,10^

### Statistical Analysis

We used descriptive statistics to characterize all patients who were tested and hospitalized during the study and follow up periods and who had recorded values for specified variables. Age was expressed as a median and interquartile range, with ranges provided that align with current citywide reporting. Categorical variables were summarized with counts and percentages.

All analyses were performed with the use of R software version 3.6.2 (R Foundation for Statistical Computing). Tests of significance were conducted using Pearson chi-square tests. We considered a p-value of <0.05 to be statistically significant.

This study was approved by the Biomedical Research Alliance of New York Institutional Review Board. Waivers of informed consent and of the Health Information Portability and Privacy Act were granted due to the retrospective nature of the study.

## Results

During the study period, 22254 patients were tested for COVID-19. 78 (0.3%) of these patients did not have a test result available at the end of the follow up period and were excluded from this analysis. Of the 22176 patients with a test result, 13442 (61%) tested positive; 6248 (46%) of patients who tested positive were hospitalized and 1724 (28%) of hospitalized patients died (**Figure 1**).

**Figure 1:**
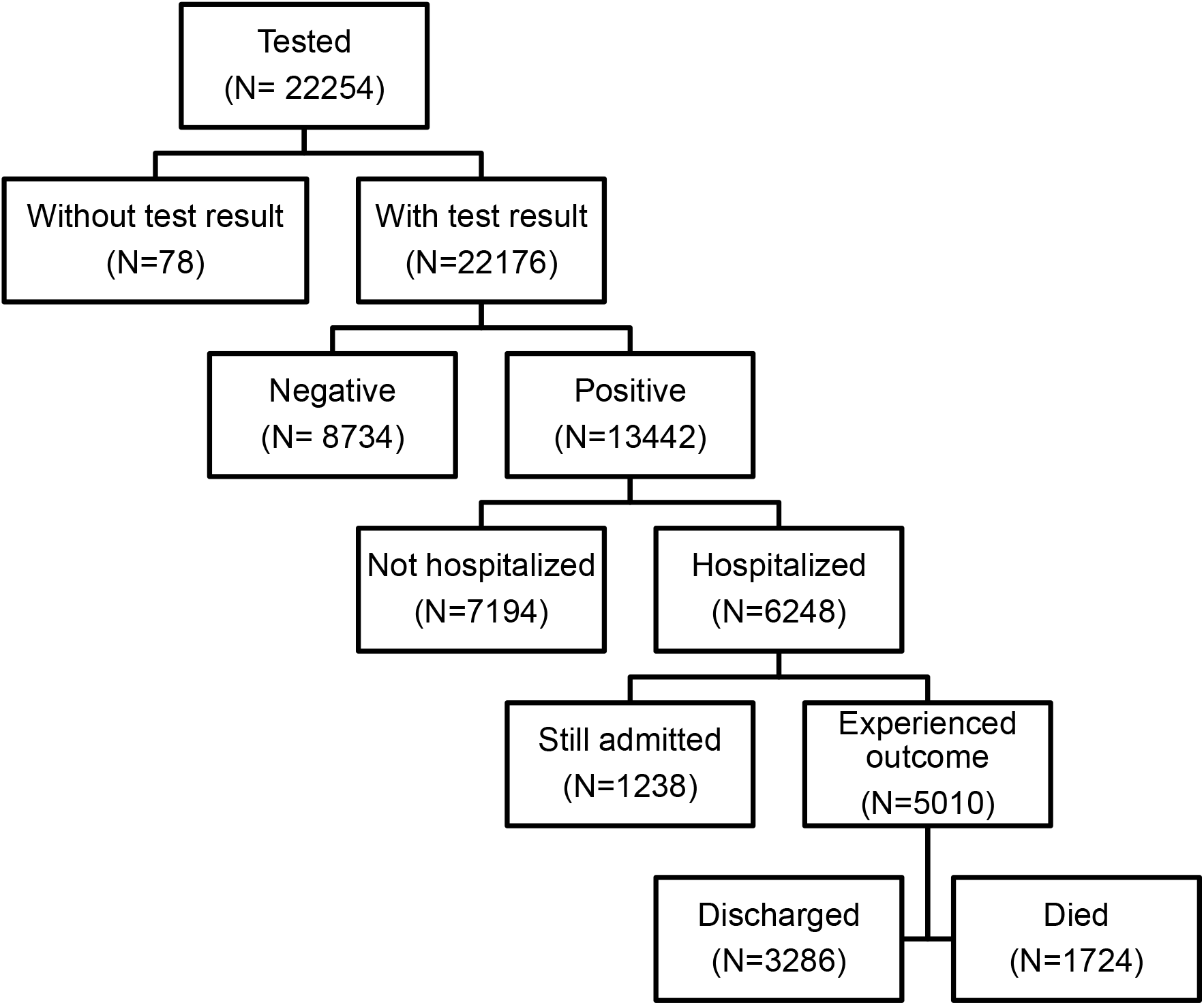
Flow Diagram of Included Patients.

Of the 13442 individuals with laboratory-confirmed SARS-CoV-2 infection, 7481 (56%) were male; males were significantly more likely to test positive than females (56% vs. 44% of all positive tests, respectively, p<0.001). The median age of individuals who tested positive was 52.7 (39.5, 64.5). 5506 (41%) patients were 45-64 years old and 104 (1%) were under 18 years of age. Older age was significantly associated with a positive test result (21% among ages 0-17 and 78% among age 75 and older, p<0.001).

Race/ethnicity was available for 22107 (99.7%) of the 22176 patients with a test result. Of those who tested positive, 3518 (26%) were Black and 4593 (34%) were Hispanic. An additional 3276 (24%) were of other or unknown race/ethnicity. Black race and Hispanic ethnicity were significantly associated with testing positive. See **Table 1** for full results and **Figure 2** for test results by demographic groups.

**Table 1:** Characteristics and Outcomes of Tested Patients.

|  |  | Patients by Test Result (No., %) |  |  |  |
| --- | --- | --- | --- | --- | --- |
|  |  | All | Negative | Positive | P value<sup>a</sup> |
|  | Total | 22176 (100) | 8734 (39) | 13442 (61) | --- |
| Sex | Female | 10965 (49) | 5008 (57) | 5957 (44) | *Reference* |
|  | Male | 11207 (51) | 3726 (43) | 7481 (56) | <0.001 |
|  | Unknown/declined | 4 (0) | 0 (0) | 4 (0) | --- |
| Age | Median (IQR) | 50.2 (36.6, 61.9) | 45.6 (33.2, 58.0) | 52.7 (39.5, 64.5) | --- |
|  | 0-17 | 491 (2) | 387 (4) | 104 (1) | <0.001 |
|  | 18-44 | 8472 (38) | 3886 (44) | 4586 (34) | *Reference* |
|  | 45-64 | 8847 (40) | 3341 (38) | 5506 (41) | <0.001 |
|  | 65-74 | 2441 (11) | 687 (8) | 1754 (13) | <0.001 |
|  | 75+ | 1925 (9) | 433 (5) | 1492 (11) | <0.001 |
| Race/ Ethnicity<sup>b</sup> | Black | 5790 (26) | 2272 (26) | 3518 (26) | <0.001 |
|  | Hispanic | 6249 (28) | 1656 (19) | 4593 (34) | <0.001 |
|  | White | 2316 (10) | 1151 (13) | 1165 (9) | *Reference* |
|  | Asian or Pacific Islander | 1739 (8) | 876 (10) | 863 (6) | 0.400 |
|  | Other | 6013 (27) | 2737 (31) | 3276 (24) | 0.002 |
| BMI<sup>c</sup> | BMI <25 | 6338 (40) | 1652 (26) | 2196 (35) | *Reference* |
|  | BMI 25 to <30 | 4613 (29) | 1473 (32) | 3127 (68) | 0.004 |
|  | BMI 30 to < 40 | 3973 (25) | 1182 (30) | 2784 (70) | <0.001 |
|  | BMI ≥40 | 904 (6) | 250 (28) | 652 (72) | 0.002 |
| Comorbidities<sup>d</sup> | Diabetes | 3752 (23) | 1042 (17) | 2710 (27) | <0.001 |
|  | Hypertension | 4424 (27) | 1381 (22) | 3043 (30) | <0.001 |
|  | Arrhythmia | 998 (6) | 364 (6) | 634 (6) | 0.299 |
|  | CVD | 3291 (20) | 1205 (19) | 2086 (21) | 0.057 |
|  | CHF | 840 (5) | 319 (5) | 521 (5) | 0.984 |
|  | Asthma | 1421 (9) | 711 (11) | 710 (7) | <0.001 |
|  | COPD | 583 (4) | 299 (5) | 284 (3) | <0.001 |
|  | Liver disease | 461 (3) | 188 (3) | 273 (3) | 0.243 |
|  | Chronic kidney disease | 1129 (7) | 320 (5) | 809 (8) | <0.001 |
|  | Cancer | 1554 (9) | 557 (9) | 997 (10) | 0.061 |
|  | HIV | 277 (2) | 118 (2) | 159 (2) | 0.132 |
|  | ≥1 chronic disease<sup>e</sup> | 7072 (43) | 2403 (38) | 4669 (46) | <0.001 |

<sup>a</sup> P values for sex, age, race/ethnicity, and BMI are from pairwise $\chi^2$ tests with female sex, ages 18-44, White, and BMI <25 as reference categories, respectively. P values for comorbidities are from pairwise $\chi^2$ tests with the group without the comorbidity (not presented in the table) as the reference category.
<sup>b</sup> Race/ethnicity was available for 22107 (99.7%) patients.
<sup>c</sup> BMI was available for 15828 (71%) patients.
<sup>d</sup> Comorbidities were available for 16420 (74%) patients.
<sup>e</sup> Flag for $\geq 1$ chronic disease includes diabetes, hypertension, cardiovascular disease, asthma, chronic obstructive pulmonary disorder, and chronic kidney disease.

**Figure 2:**
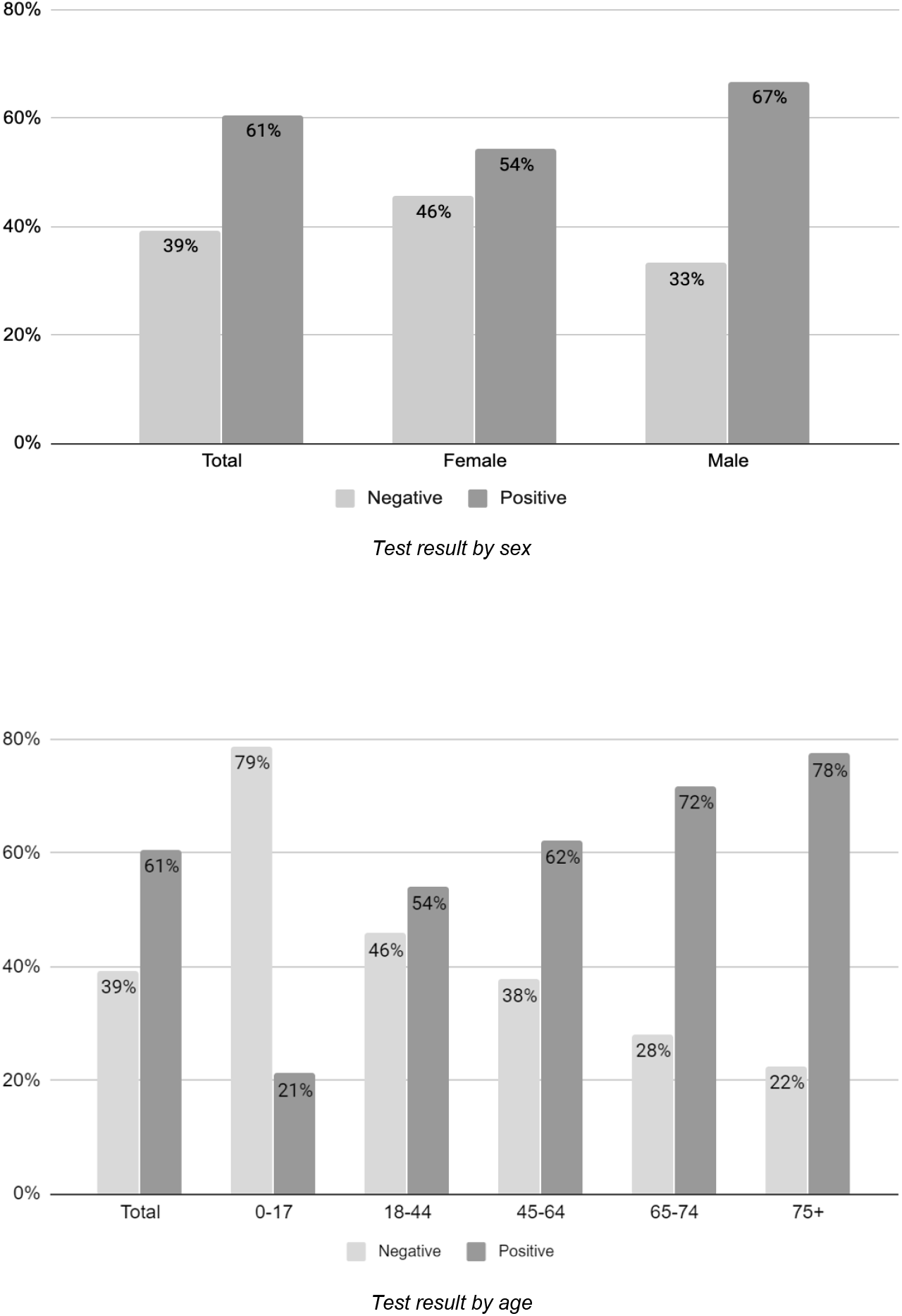
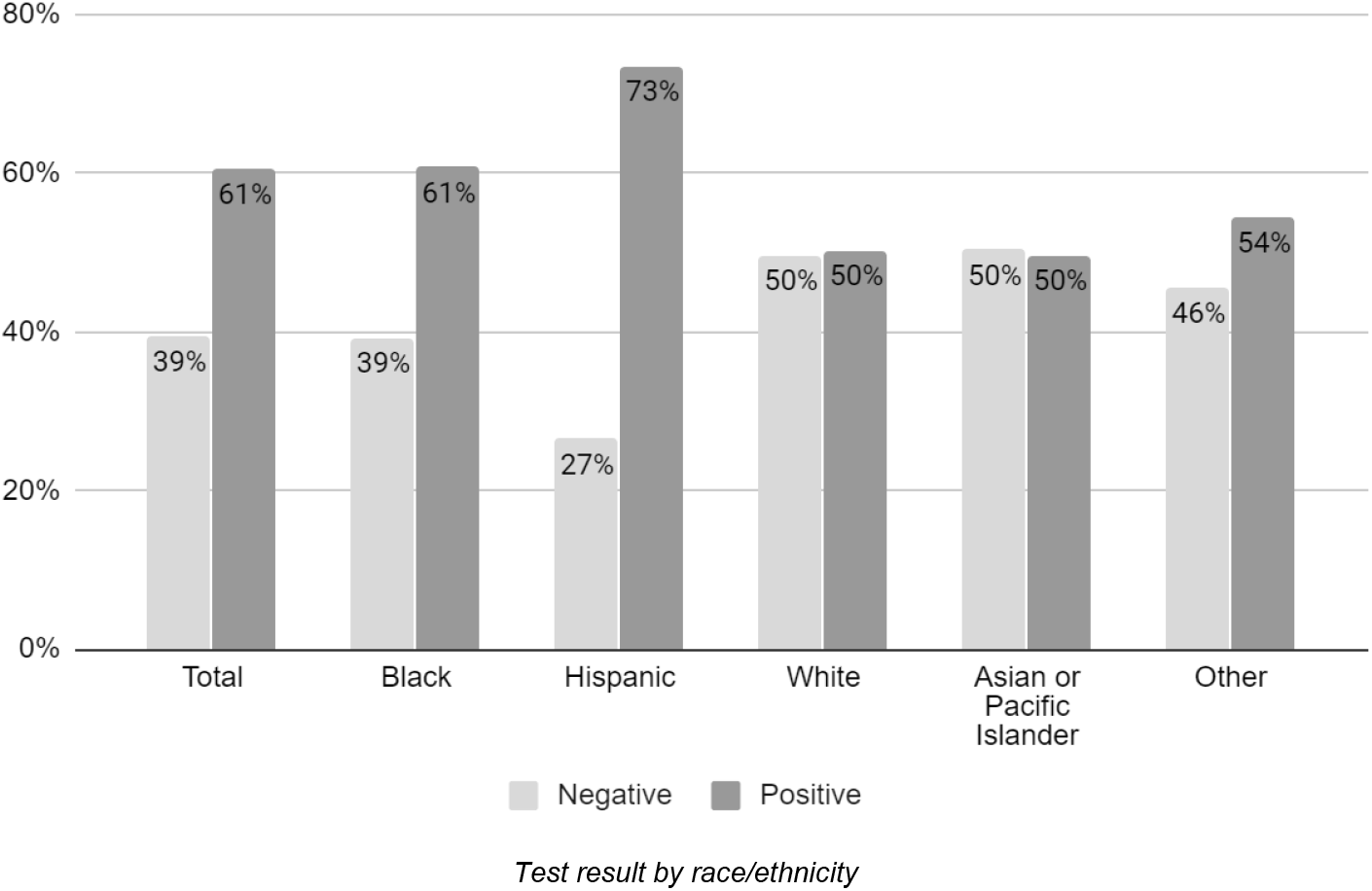
Test Results by Demographic Factors.

BMI was available for 15828 (71%) of tested individuals; we found that BMI ≥25 was significantly associated with testing positive. A diagnosis history was available for 16420 (74%) individuals who were tested. The most common chronic diseases were diabetes (27%), HTN (30%), and CVD (21%). The presence of at least one chronic disease was significantly associated with testing positive (46% vs. 38%), particularly for those with diabetes (27% vs 17%), HTN (30% vs. 22%), and CKD (8% vs. 5%) (for all, p<0.001). Individuals with asthma and COPD were significantly less likely to test positive (7% vs. 11% and 3% vs. 5%, respectively, p<0.001 for both) (**Table 1**).

**Table 2** displays the demographics and clinical characteristics and hospitalization outcome of patients with a positive test result. Nearly half of patients who tested positive (6248, 46%) were hospitalized. Males comprised nearly two-thirds (3851, 62%) of hospitalized patients, and male sex was significantly associated with hospitalization (p<0.001). As compared to females, a greater proportion of males were hospitalized (51% vs. 40%).

**Table 2:** Characteristics and Outcomes of Patients Testing Positive.

|  |  | Patients who Tested Positive by Outcome (No., %) |  |  |  |
| --- | --- | --- | --- | --- | --- |
|  |  | Total | Not Hospitalized | Hospitalized | P value<sup>a</sup> |
|  | Total | 13442 (100) | 7194 (54) | 6248 (46) | --- |
| Sex | Female | 5957 (44) | 3561 (49) | 2396 (38) | *Reference* |
|  | Male | 7481 (56) | 3630 (50) | 3851 (62) | <0.001 |
|  | Unknown / declined | 4 (0) | 3 (0) | 1 (0) | --- |
| Age | *Median (IQR)* | 52.7 (39.5, 64.5) | 45.2 (34.5, 56.1) | 61.6 (49.7, 72.9) | --- |
|  | 0-17 | 104 (1) | 72 (1) | 32 (1) | 0.138 |
|  | 18-44 | 4586 (34) | 3486 (48) | 1100 (18) | *Reference* |
|  | 45-64 | 5506 (41) | 3015 (42) | 2491 (40) | <0.001 |
|  | 65-74 | 1754 (13) | 451 (6) | 1303 (21) | <0.001 |
|  | 75+ | 1492 (11) | 170 (2) | 1322 (21) | <0.001 |
| Race/ Ethnicity<sup>b</sup> | Black | 3518 (26) | 1568 (22) | 1950 (31) | <0.001 |
|  | Hispanic | 4593 (34) | 2491 (35) | 2102 (34) | 0.99 |
|  | White | 1165 (9) | 631 (9) | 534 (9) | *Reference* |
|  | Asian or Pacific Islander | 863 (6) | 489 (7) | 374 (6) | 0.28 |
|  | Other | 3276 (24) | 1988 (28) | 1288 (21) | <0.001 |
| BMI<sup>c</sup> | BMI <25 | 2196 (25) | 769 (25) | 1427 (25) | *Reference* |
|  | BMI 25 to <30 | 3127 (36) | 1173 (38) | 1954 (35) | 0.069 |
|  | BMI 30 to < 40 | 2784 (32) | 1009 (33) | 1775 (31) | 0.386 |
|  | BMI >40 | 652 (7) | 149 (5) | 503 (9) | <.001 |
| Comorbidities<sup>d</sup> | Diabetes | 2710 (27) | 665 (17) | 2045 (33) | <0.001 |
|  | Hypertension | 3043 (30) | 756 (19) | 2287 (37) | <0.001 |
|  | Arrhythmia | 634 (6) | 115 (3) | 519 (8) | <0.001 |
|  | CVD | 2086 (21) | 591 (15) | 1495 (24) | <0.001 |
|  | CHF | 521 (5) | 58 (1) | 463 (7) | <0.001 |
|  | Asthma | 710 (7) | 283 (7) | 427 (7) | 0.668 |
|  | COPD | 284 (3) | 51 (1) | 233 (4) | <0.001 |
|  | Liver disease | 273 (3) | 83 (2) | 190 (3) | 0.004 |
|  | Chronic kidney disease | 809 (8) | 103 (3) | 706 (11) | <0.001 |
|  | Cancer | 997 (10) | 396 (10) | 601 (10) | 0.663 |
|  | HIV | 159 (2) | 65 (2) | 94 (2) | 0.689 |
|  | >1 chronic disease<sup>e</sup> | 4669 (46) | 1400 (35) | 3269 (53) | <0.001 |

<sup>a</sup> P values for sex, age, race/ethnicity, and BMI are from pairwise $\chi^2$ tests with female sex, ages 18-44, White, and BMI <25 as reference categories, respectively. P values for comorbidities are from pairwise $\chi^2$ tests with the group without the comorbidity (not presented in the table) as the reference category.
<sup>b</sup> Race/ethnicity was available for 13415 (99.8%) patients.
<sup>c</sup> BMI was available for 8759 (65%) patients.
<sup>d</sup> Comorbidities were available for 10169 (76%) patients.
<sup>e</sup> Flag for $\geq 1$ chronic disease includes diabetes, hypertension, cardiovascular disease, asthma, chronic obstructive pulmonary disorder, and chronic kidney disease.

The median age of hospitalized patients was 61.6 years (IQR 49.7, 72.9); the overwhelming majority of patients were over the age of 45, with 2491 (40%) between ages 45 and 64, 1303 (21%) between ages 65 and 74, and an additional 1322 (21%) age 75 and older. Among individuals who tested positive, older age was significantly associated with hospitalization, with 24% of individuals age 18-44 and 89% of individuals age 75 and older hospitalized.

Blacks and Hispanics each comprised approximately one-third of hospitalized patients (31% and 34%, respectively); black race was significantly associated with hospitalization. Additionally, Blacks were more likely to be hospitalized than individuals of other racial/ethnic groups (55% vs. 46% for both Hispanics and Whites and 43% for APIs). Full results of hospitalization rates by demographic factors are in **Figure 3**.

**Figure 3:**
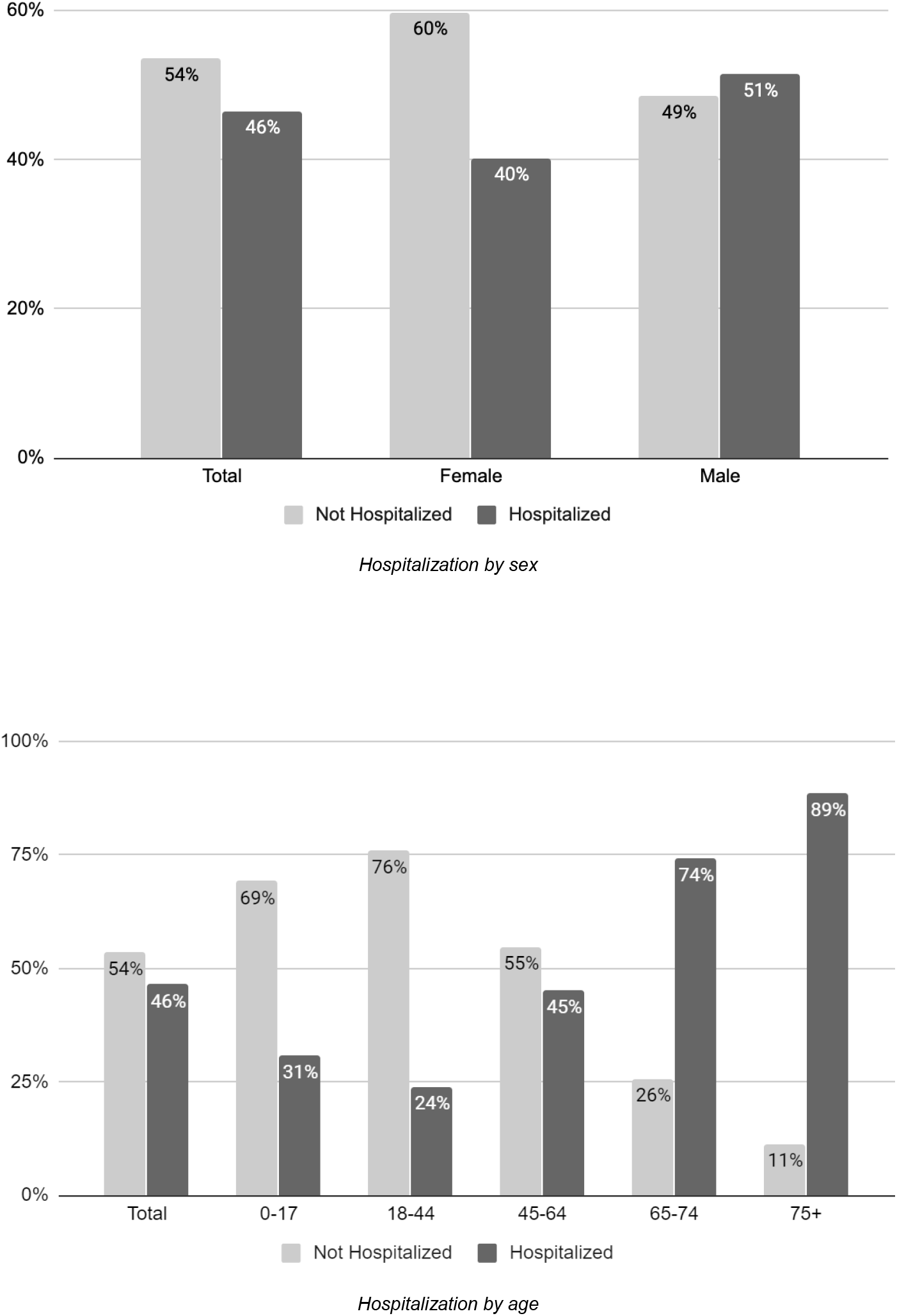
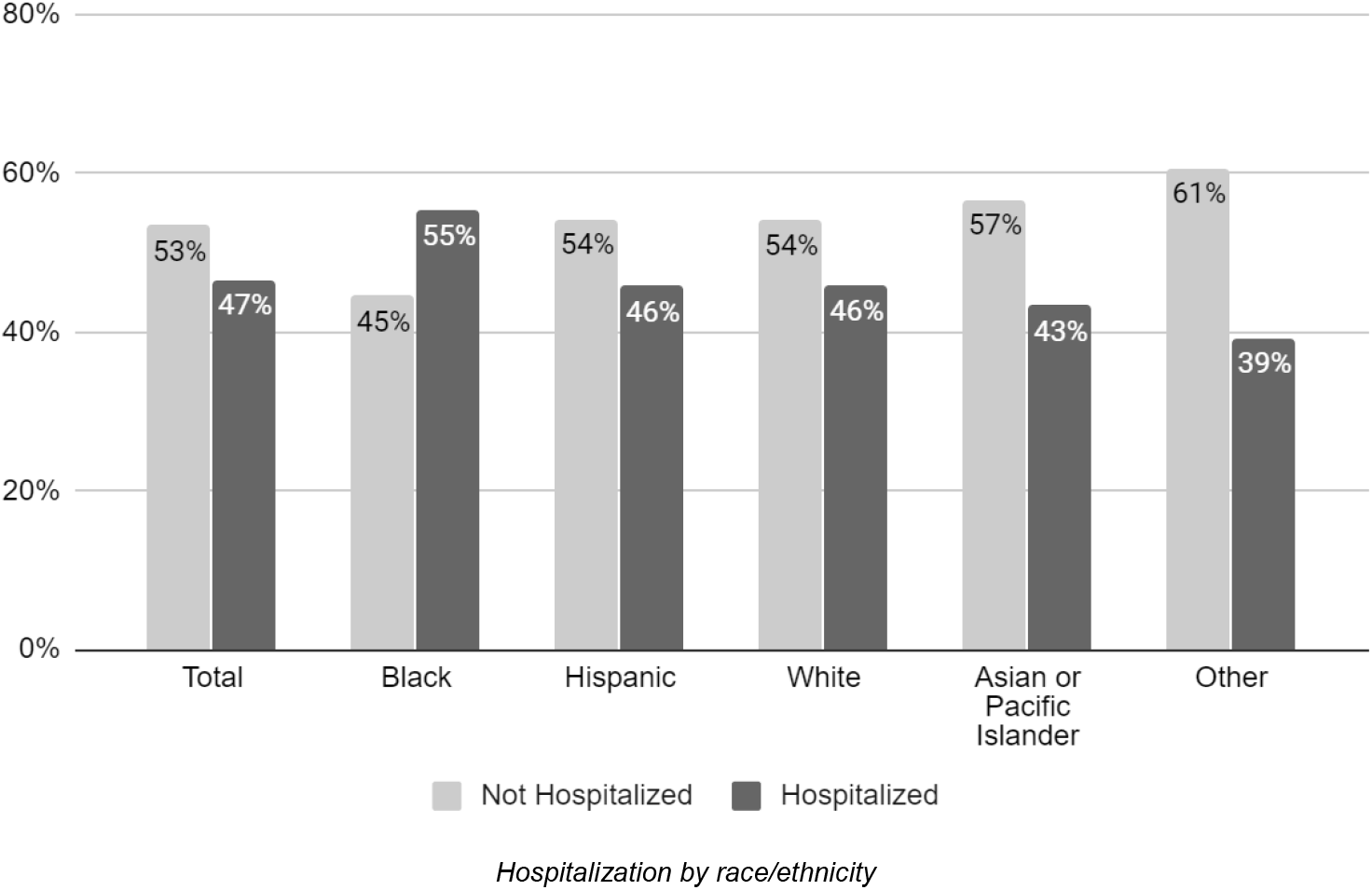
Hospitalization by Demographic Factors.

Among individuals who tested positive with a recorded BMI (8759; 65%), hospitalization was significantly more likely among individuals with BMI ≥40 (9% vs. 5%). A diagnosis history was available for 10169 (76%) of patients who tested positive. Chronic diseases were common among patients who were hospitalized, with 53% having one or more chronic disease, including diabetes (33%), HTN (37%), CVD (24%), and chronic kidney disease (11%). Having at least one chronic disease was significantly associated with hospitalization (53% vs. 35%) as were diabetes (33% vs. 17%), HTN (37% vs. 19%), arrhythmia (8% vs. 3%), CVD (24% vs. 15%), CHF (7% vs. 1%), COPD (4% vs. 1%), liver disease (3% vs. 2%), and CKD (11% vs. 3%) (for all, p<0.001). We found no significant association between asthma, cancer, or HIV and hospitalization (**Table 2**).

**Table 3** displays the demographics and clinical characteristics and clinical outcome of hospitalized patients. Of the 6248 hospitalized patients, 1238 (20%) were still admitted at the end of the study period, 3286 (53%) were discharged, and 1724 (28%) died.

**Table 3:** Characteristics and Outcomes of Hospitalized Patients.

|  |  | Hospitalized Patients (No., %) |  |  |  |  |
| --- | --- | --- | --- | --- | --- | --- |
|  |  | All | Still Admitted | Discharged | Died | P value<sup>a</sup> |
|  | Total | 6248 (100) | 1238 (20) | 3286 (53) | 1724 (28) | --- |
| Sex | Female | 2396 (38) | 441 (36) | 1318 (40) | 637 (37) | *Reference* |
|  | Male | 3851 (62) | 796 (64) | 1968 (60) | 1087 (63) | 0.032 |
|  | Unknown/ declined | 1 (0) | 1 (0) | 0 (0) | 0 (0) | --- |
| Age | Median (IQR) | 61 [49.7, 72.9] | 62.5 [53.6, 71.9] | 56.3 [44.2, 67.7] | 71 [60.0,80.9] | --- |
|  | 0-17 | 32 (1) | 1 (0) | 30 (1) | 1 (0) | --- |
|  | 18-44 | 1100 (18) | 146 (12) | 826 (25) | 128 (7) | *Reference* |
|  | 45-64 | 2491 (40) | 561 (45) | 1447 (44) | 483 (28) | <.001 |
|  | 65-74 | 1302 (21) | 298 (24) | 572 (17) | 433 (25) | <.001 |
|  | 75+ | 1322 (21) | 232 (19) | 411 (13) | 679 (39) | <.001 |
| Race/<br>Ethnicity | Black | 1950 (31) | 376 (30) | 1068 (33) | 506 (29) | <.001 |
|  | Hispanic | 2102 (34) | 414 (33) | 1160 (35) | 528 (31) | <.001 |
|  | White | 534 (9) | 94 (8) | 232 (7) | 208 (12) | *Reference* |
|  | Asian or Pacific Islander | 374 (6) | 78 (6) | 186 (6) | 110 (6) | 0.008 |
|  | Other | 1288 (21%) | 276 (22) | 640 (19) | 372 (22) | <.001 |
| BMI<sup>b</sup> | BMI <25 | 1427 (25) | 302 (25) | 731 (25) | 394 (26) | *Reference* |
|  | BMI 25 to <30 | 1954 (35) | 394 (33) | 1059 (36) | 501 (33) | 0.125 |
|  | BMI 30 to < 40 | 1775 (31) | 388 (33) | 945 (32) | 442 (30) | 0.104 |
|  | BMI >40 | 503 (9) | 106 (9) | 238 (8) | 159 (11) | 0.084 |
| Comorbidities<sup>c</sup> | Diabetes | 2045 (33) | 420 (34) | 1028 (32) | 597 (35) | 0.032 |
|  | Hypertension | 2287 (37) | 497 (40) | 1151 (35) | 639 (37) | 0.241 |
|  | Arrhythmia | 519 (8) | 193 (11) | 223 (7) | 193 (11) | 0.001 |
|  | CVD | 1495 (24) | 297 (24) | 722 (22) | 476 (28) | 0.001 |
|  | CHF | 463 (7) | 105 (9) | 199 (6) | 159 (9) | 0.001 |
|  | Asthma | 427 (7) | 75 (6) | 249 (8) | 103 (6) | 0.033 |
|  | COPD | 233 (4) | 43 (3) | 121 (4) | 69 (4) | 0.671 |
|  | Liver disease | 190 (3) | 44 (4) | 99 (3) | 47 (3) | 0.592 |
|  | Chronic kidney disease | 706 (11) | 153 (12) | 295 (9) | 258 (15) | 0.001 |
|  | Cancer | 601 (10) | 116 (9) | 305 (9) | 180 (10) | 0.243 |
|  | HIV | 94 (2) | 16 (1) | 58 (2) | 20 (1) | 0.119 |
|  | >1 chronic disease<sup>d</sup> | 3269 (53) | 673 (55) | 1684 (52) | 912 (53) | 0.442 |

<sup>a</sup> P values for sex, age, race/ethnicity, and BMI are from pairwise $\chi^2$ tests with female sex, ages 18-44, White, and BMI <25 as reference categories, respectively. P values for comorbidities are from pairwise $\chi^2$ tests with the group without the comorbidity (not presented in the table) as the reference category.
<sup>b</sup> BMI was available for 5659 (91%) patients.
<sup>c</sup> Comorbidities were available for 6200 (99%) patients.
<sup>e</sup> Flag for $\geq 1$ chronic disease includes diabetes, hypertension, cardiovascular disease, asthma, chronic obstructive pulmonary disorder, and chronic kidney disease.

Males comprised nearly two-thirds (1087, 63%) of patients who died, and male sex was significantly associated with death. The median age of patients who died was 71.0 years (IQR 60.0, 80.9); the majority of patients were over the age of 65, with 433 (25%) between ages 65 and 74 and 679 (39%) age 75 and older. Increasing age was significantly associated with death.

Blacks and Hispanics each comprised approximately one-third of all patients who died (29% and 31%, respectively), and Whites comprised 12%. White race was significantly associated with death, with a greater proportion of Whites dying than among all other racial/ethnic groups (47% vs. 32% for Blacks, 31% for Hispanics, and 37% for APIs). Full results of death rates by demographic factors are in **Figure 4**.

**Figure 4:**
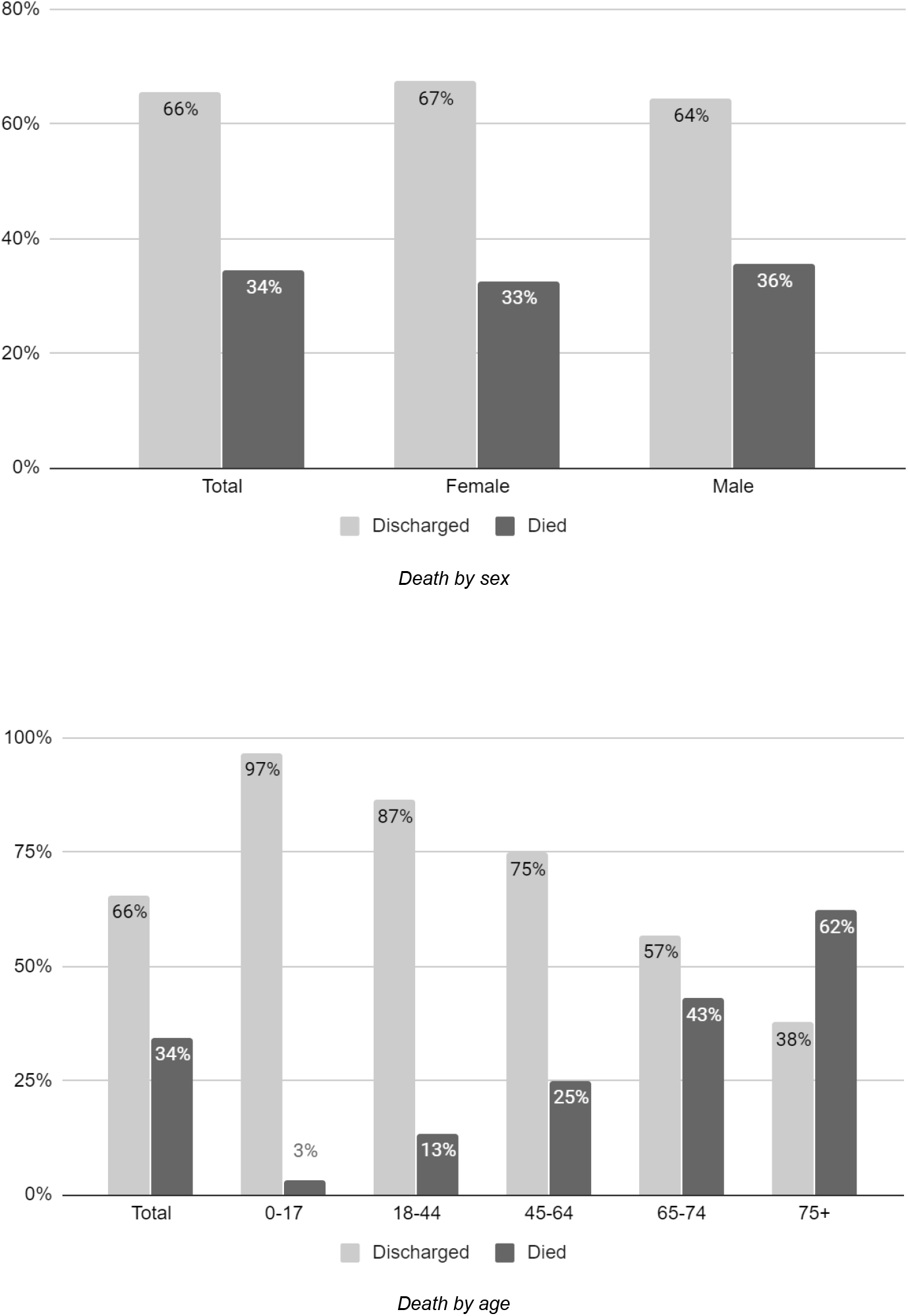
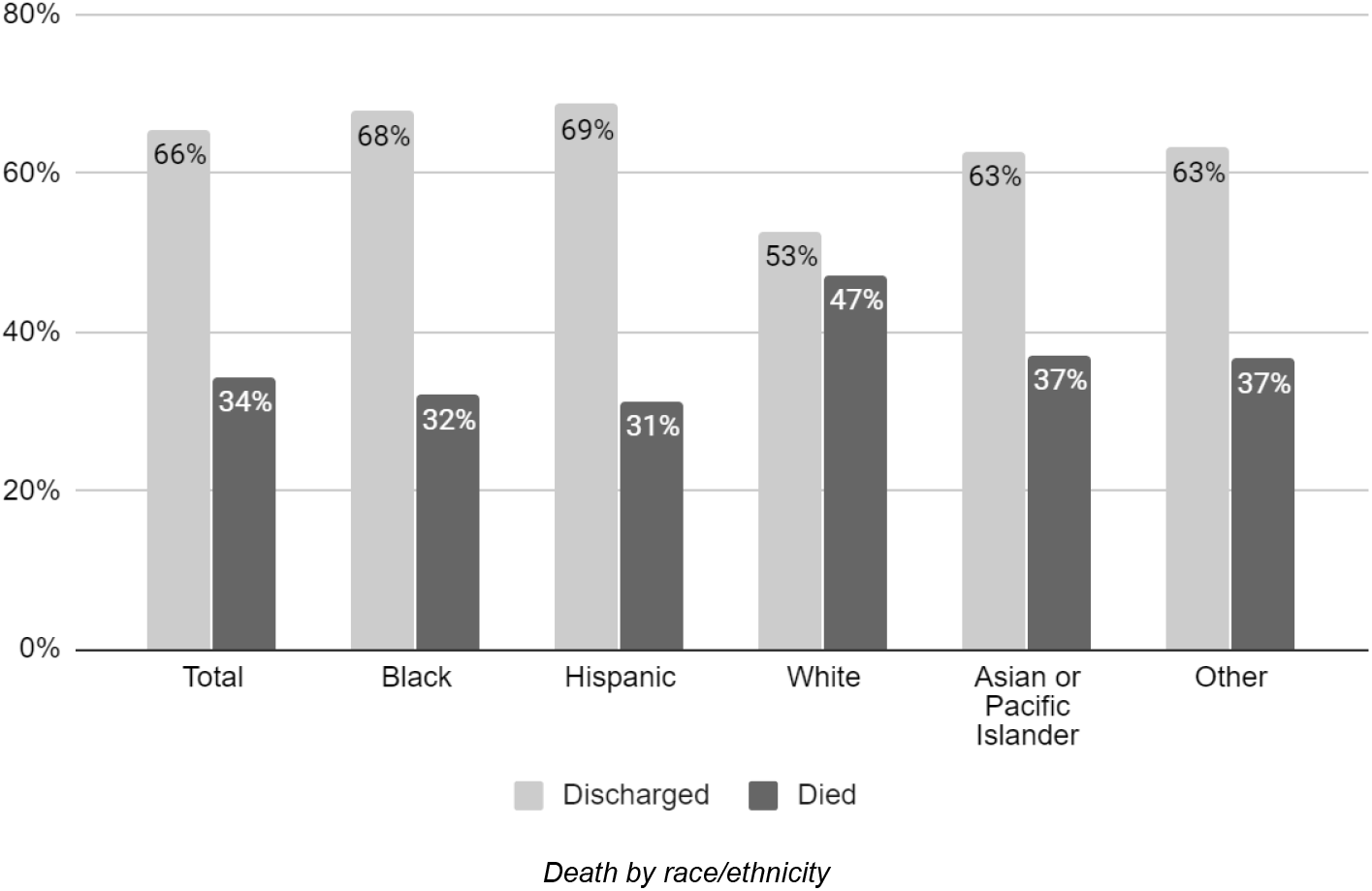
Death by Demographic Factors.

Among the 5659 (90%) hospitalized patients with a BMI value, individuals with BMI ≥40 were more likely to die than be discharged (11% vs. 8%), but this association was not significant (p=0.084). Chronic diseases were common among patients who died, with 53% having one or more chronic disease, including diabetes (35%), HTN (37%), CVD (28%), CHF (9%), and CKD (15%). Diabetes, arrhythmia, CVD, CHF, and CKD were significantly associated with death (p<0.001), whereas asthma was significantly associated with discharged (p=0.033) (**Table 3**).

We observed consistent differences across racial/ethnic groups in the age distribution across all outcomes, with Hispanics younger and Whites older than other racial/ethnic groups (**Table 4**). We also observed differences by race/ethnicity in the prevalence of comorbidities among patients who were hospitalized and died (**Table 5**). Among patients who were hospitalized, Hispanics and Blacks had a higher prevalence of obesity than other groups, with 13% of Blacks having a BMI ≥40 as compared to 6% of Hispanics and 8% of Whites. Hospitalized Blacks were also more likely to have one or more chronic disease than other racial/ethnic groups (64% of Blacks vs. 50% of Hispanics and 52% of Whites) and also had a higher prevalence of diabetes, HTN, CVD, CHF, and CKD. These disparities were largely consistent among patients who died, with 15% of Blacks having a BMI ≥40 as compared to 7% of Hispanics and 8% of Whites and the highest prevalence of one or more chronic diseases among all racial/ethnic groups (62% vs 54% for both Hispanics and Whites and 51% for APIs), including diabetes, HTN, and CKD. Whites who died had the highest prevalence of arrhythmia (17%), CVD (36%), and CHF (15%), and COPD (7%) of all racial/ethnic groups.

**Table 4:** Age Distribution of Patients by Race/Ethnicity.

|  | **Total** | **Black** | **Hispanic** | **White** | **Asian / Pacific**<br>**Islander** | **Other** |
| --- | --- | --- | --- | --- | --- | --- |
| **Tested Positive (No., %)** |  |  |  |  |  |  |
| 0-17 | 103 (1) | 18 (1) | 49 (1) | 5 (0) | 6 (1) | 25 (1) |
| 18-44 | 4577 (34) | 845 (24) | 1732 (38) | 340 (29) | 291 (34) | 1369 (42) |
| 45-64 | 5493 (41) | 1576 (45) | 1918 (42) | 417 (36) | 350 (41) | 1232 (38) |
| 65-74 | 1752 (13) | 574 (16) | 489 (11) | 180 (15) | 139 (16) | 370 (11) |
| ≥75 | 1490 (11) | 505 (14) | 405 (9) | 223 (19) | 77 (9) | 280 (9) |
| **Total** | **13415** | **3518** | **4593** | **1165** | **863** | **3276** |
| **Hospitalized (No., %)** |  |  |  |  |  |  |
| 0-17 | 32 (1) | 4 (0) | 17 (1) | 1 (0) | 1 (0) | 9 (1) |
| 18-44 | 1100 (18) | 219 (11) | 514 (24) | 46 (9) | 55 (15) | 266 (21) |
| 45-64 | 2491 (40) | 814 (42) | 873 (42) | 148 (28) | 140 (37) | 516 (40) |
| 65-74 | 1303 (21) | 462 (24) | 347 (17) | 126 (24) | 107 (29) | 261 (20) |
| ≥75 | 1322 (21) | 451 (23) | 351 (17) | 213 (40) | 71 (19) | 236 (18) |
| **Total** | **6248** | **1950** | **2102** | **534** | **374** | **1288** |
| **Died (No., %)** |  |  |  |  |  |  |
| 0-17 | 1 (0) | 0 (0) | 1 (0) | 0 (0) | 0 (0) | 0 (0) |
| 18-44 | 128 (7) | 25 (5) | 59 (11) | 5 (2) | 4 (4) | 34 (9) |
| 45-64 | 483 (28) | 138 (27) | 168 (32) | 35 (17) | 27 (25) | 115 (31) |
| 65-74 | 433 (25) | 13 (26) | 115 (22) | 42 (20) | 37 (34) | 106 (28) |
| ≥75 | 679 (39) | 209 (41) | 185 (35) | 126 (61) | 42 (38) | 117 (31) |
| **Total** | **1724** | **506** | **528** | **208** | **110** | **372** |

**Table 5:**
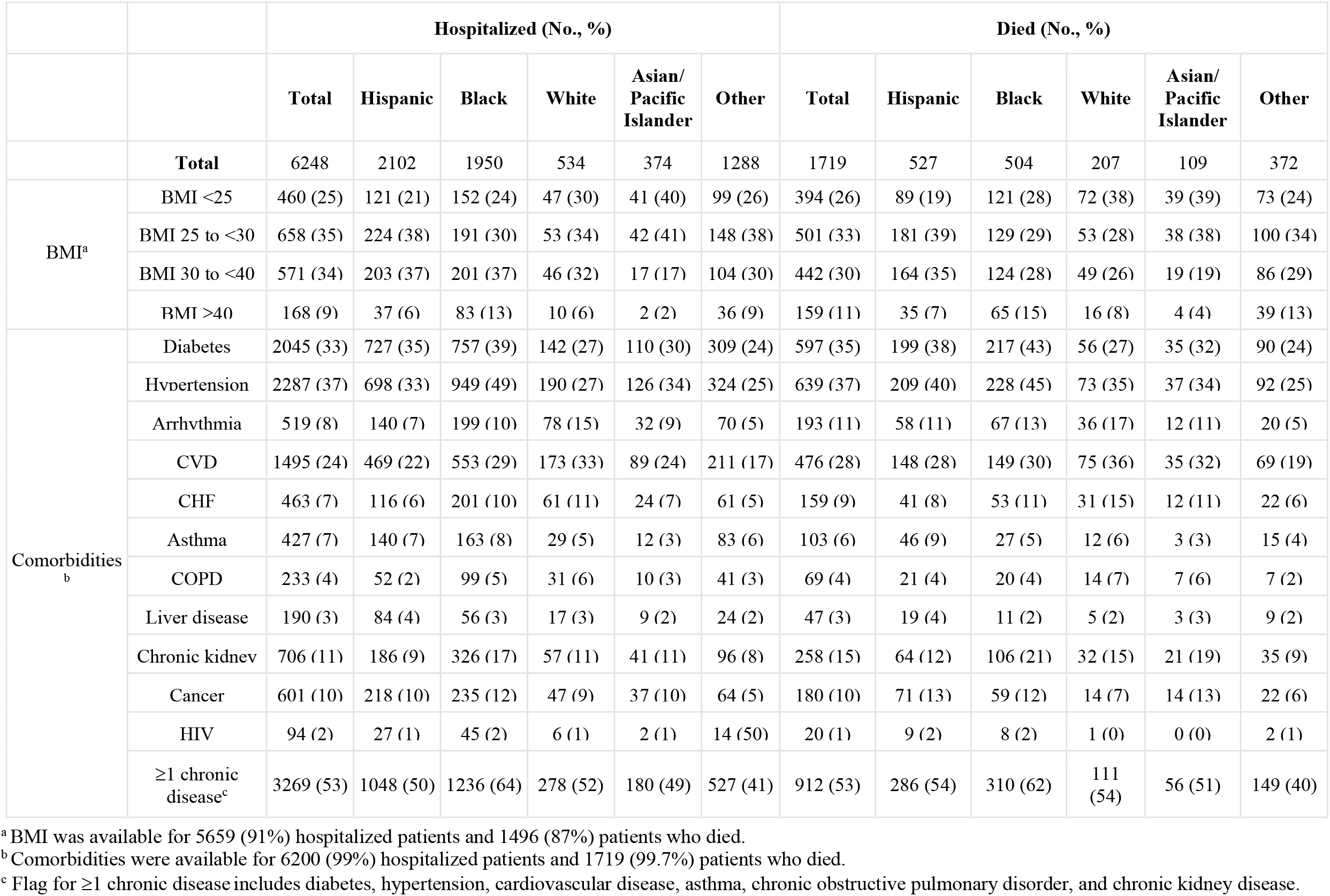
Comorbidities of Patients by Race/Ethnicity.

|  |  | Hospitalized (No., %) |  |  |  |  |  | Died (No., %) |  |  |  |  |  |
| --- | --- | --- | --- | --- | --- | --- | --- | --- | --- | --- | --- | --- | --- |
|  |  | Total | Hispanic | Black | White | Asian/<br>Pacific<br>Islander | Other | Total | Hispanic | Black | White | Asian/<br>Pacific<br>Islander | Other |
|  | **Total** | 6248 | 2102 | 1950 | 534 | 374 | 1288 | 1719 | 527 | 504 | 207 | 109 | 372 |
| BMI<sup>a</sup> | BMI <25 | 460 (25) | 121 (21) | 152 (24) | 47 (30) | 41 (40) | 99 (26) | 394 (26) | 89 (19) | 121 (28) | 72 (38) | 39 (39) | 73 (24) |
|  | BMI 25 to <30 | 658 (35) | 224 (38) | 191 (30) | 53 (34) | 42 (41) | 148 (38) | 501 (33) | 181 (39) | 129 (29) | 53 (28) | 38 (38) | 100 (34) |
|  | BMI 30 to <40 | 571 (34) | 203 (37) | 201 (37) | 46 (32) | 17 (17) | 104 (30) | 442 (30) | 164 (35) | 124 (28) | 49 (26) | 19 (19) | 86 (29) |
|  | BMI ≥40 | 168 (9) | 37 (6) | 83 (13) | 10 (6) | 2 (2) | 36 (9) | 159 (11) | 35 (7) | 65 (15) | 16 (8) | 4 (4) | 39 (13) |
| Comorbidities<sup>b</sup> | Diabetes | 2045 (33) | 727 (35) | 757 (39) | 142 (27) | 110 (30) | 309 (24) | 597 (35) | 199 (38) | 217 (43) | 56 (27) | 35 (32) | 90 (24) |
|  | Hypertension | 2287 (37) | 698 (33) | 949 (49) | 190 (27) | 126 (34) | 324 (25) | 639 (37) | 209 (40) | 228 (45) | 73 (35) | 37 (34) | 92 (25) |
|  | Arrhythmia | 519 (8) | 140 (7) | 199 (10) | 78 (15) | 32 (9) | 70 (5) | 193 (11) | 58 (11) | 67 (13) | 36 (17) | 12 (11) | 20 (5) |
|  | CVD | 1495 (24) | 469 (22) | 553 (29) | 173 (33) | 89 (24) | 211 (17) | 476 (28) | 148 (28) | 149 (30) | 75 (36) | 35 (32) | 69 (19) |
|  | CHF | 463 (7) | 116 (6) | 201 (10) | 61 (11) | 24 (7) | 61 (5) | 159 (9) | 41 (8) | 53 (11) | 31 (15) | 12 (11) | 22 (6) |
|  | Asthma | 427 (7) | 140 (7) | 163 (8) | 29 (5) | 12 (3) | 83 (6) | 103 (6) | 46 (9) | 27 (5) | 12 (6) | 3 (3) | 15 (4) |
|  | COPD | 233 (4) | 52 (2) | 99 (5) | 31 (6) | 10 (3) | 41 (3) | 69 (4) | 21 (4) | 20 (4) | 14 (7) | 7 (6) | 7 (2) |
|  | Liver disease | 190 (3) | 84 (4) | 56 (3) | 17 (3) | 9 (2) | 24 (2) | 47 (3) | 19 (4) | 11 (2) | 5 (2) | 3 (3) | 9 (2) |
|  | Chronic kidney | 706 (11) | 186 (9) | 326 (17) | 57 (11) | 41 (11) | 96 (8) | 258 (15) | 64 (12) | 106 (21) | 32 (15) | 21 (19) | 35 (9) |
|  | Cancer | 601 (10) | 218 (10) | 235 (12) | 47 (9) | 37 (10) | 64 (5) | 180 (10) | 71 (13) | 59 (12) | 14 (7) | 14 (13) | 22 (6) |
|  | HIV | 94 (2) | 27 (1) | 45 (2) | 6 (1) | 2 (1) | 14 (50) | 20 (1) | 9 (2) | 8 (2) | 1 (0) | 0 (0) | 2 (1) |
|  | ≥1 chronic disease<sup>c</sup> | 3269 (53) | 1048 (50) | 1236 (64) | 278 (52) | 180 (49) | 527 (41) | 912 (53) | 286 (54) | 310 (62) | 111 (54) | 56 (51) | 149 (40) |

<sup>a</sup> BMI was available for 5659 (91%) hospitalized patients and 1496 (87%) patients who died.
<sup>b</sup> Comorbidities were available for 6200 (99%) hospitalized patients and 1719 (99.7%) patients who died.
<sup>c</sup> Flag for $\geq 1$ chronic disease includes diabetes, hypertension, cardiovascular disease, asthma, chronic obstructive pulmonary disorder, and chronic kidney disease.

## Discussion

In this report, we describe demographic and clinical characteristics and outcomes of patients tested for COVID-19 at the largest public hospital system in the nation. Of the 22254 patients who were tested, 13442 patients tested positive, 6248 required hospitalization, and 1724 died. We found that male sex, older age, and certain chronic diseases were significantly associated with all outcomes, with racial/ethnic disparities across all outcomes.

Although male sex was significantly associated with all outcomes, we observed that the proportion of males and females who died were relatively similar (36% and 33%, respectively), unlike with testing positive and hospitalization.

Whereas BMI ≥25 was found to be associated with testing positive, only BMI ≥40 was associated with hospitalization and death, which is consistent with other reports.^3,11,12^ In addition to the comorbidities associated with a positive test result, cardiac history and COPD were significantly associated with hospitalization. These findings are similar to previous reports showing high rates of chronic diseases, including diabetes and HTN, among COVID-19 patients.^5,8,13^ We observed fewer comorbidities associated with death, with only diabetes, CVD, CHF, arrhythmia, and CKD having a significant association; these may be driven in part by the older age of patients who died.

Although the presence of certain chronic diseases was associated with all outcomes, we observed a lower likelihood of infection among individuals with asthma and COPD and a lower likelihood of death among individuals with asthma, which is aligned with recent reports of cases in New York State and globally.^14,15^

A growing body of evidence shows a disproportionate burden of COVID-19 among Hispanics and Blacks in NYC.^4,6,7,16^ Across all outcomes, we found that Blacks and Hispanics each comprised approximately one-third of the total population, mirroring citywide rates.^17^ However, these groups were overrepresented as compared to the overall NYC population.^18^

Hispanics were more likely to test positive than any other racial/ethnic group but, once they tested positive, they were not more likely to be hospitalized or die than other groups, which is likely due to their younger age. Across all outcomes, Hispanics were younger than other racial/ethnic groups: among Hispanics only, individuals ages 18 to 64 comprised 80% of those testing positive, 66% of those hospitalized, and 44% of those who died.

Similarly, Blacks were more likely to test positive and be hospitalized than individuals of other racial/ethnic groups. This is likely driven by their higher prevalence of comorbidities, contributing to their increased risk of hospitalization.^19^ However, once hospitalized, Black race and Hispanic ethnicity were significantly associated with being discharged, while Whites were more likely to die. Our analyses of the age distribution and prevalence of comorbidities within each racial/ethnic group suggest that this is driven by the older age of and higher prevalence of CVD, CHF, and arrhythmia among Whites in our cohort. Additionally, Blacks were younger than Whites across all outcomes, indicating a disproportionate burden of illness and mortality among this group. Analyses that utilize multivariable models; control for age, demographics and comorbidities; and incorporate factors such as ICU admission and end of life decisions are necessary to better understand these disparities.

The disproportionate burden of COVID-19 among Hispanics and Blacks also may be partially explained by their overrepresentation in essential roles that require in-person work, resulting in frequent and prolonged exposure.^20^ Recent data show that only 16% of Hispanics and 20% of Blacks are able to work at home as compared to 30% of Whites and 39% of APIs, and only 9% of low-wage (<25th percentile) workers are able to work at home as compared to 62% of workers in the highest income quartile.^21^ Additional analyses that incorporate non-clinical data related to the social determinants of health will be valuable in expanding upon these findings.

Post-hoc, exploratory analyses indicate that Whites and APIs were more likely to have been tested in outpatient, non-emergency settings than Blacks and Hispanics, who were more often tested in the emergency department and inpatient settings. This may partially account for the lower positivity rates among Whites and APIs and higher hospitalization rates among Blacks and Hispanics, who may have been presenting with a longer duration and greater severity of illness. Analyses that assess testing location and symptoms at presentation can help elucidate these disparities and inform recommendations for seeking care.

To date, published reports on COVID-19 in the NYC area have been of smaller cohorts with less racial/ethnic diversity than at NYC H+H. The largest such case series reported to date was of 5700 hospitalized patients, of whom 23% were Black and 23% Hispanic; in contrast, Blacks and Hispanics comprised 31% and 34%, respectively, of the 6248 patients hospitalized for COVID-19 at NYC H+H, more closely reflecting the demographics of the overall NYC population.^6,22^ Given the diversity of NYC and the outsize and growing impact of COVID-19 on communities of color, it is important that reports on COVID-19 in the United States proportionally represent these populations.

### Limitations

Our study includes several limitations. BMI and clinical history were not uniformly available for the study population, with a greater proportion of non-hospitalized patients missing such a history. Having diagnoses recorded in the EHR was correlated with hospitalization, as, in many cases, a diagnosis history was completed upon admission as a part of the patient’s COVID-19 care plan.

Additionally, some individuals had incomplete documentation of comorbidities due to variations in the format of historical electronic databases. NYC H+H recently completed a transition to a single EHR platform for the entire health system, resulting in data from the previous platform not being uniformly carried over to the current platform.

Community testing was initially available at ambulatory sites and temporary appointment-only drive-through sites but was restricted starting on March 20 to individuals presenting at an emergency department with severe symptoms.^23,24,25^ Because the criteria for testing in NYC became more strict toward the end of the study period, individuals whose tests and hospital admissions were in the later portion of the study period were more likely to test positive or have more severe illness that required hospitalization. While this may reduce the generalizability of findings to other localities with a lower burden of COVID-19 and, accordingly, less restrictive testing policies, our findings are aligned with those from other health systems in the NYC area.

Our set of hospitalized patients includes individuals who were admitted for any reason after receiving a COVID-19 test. At the beginning of the study period, documentation in the HER for a COVID-19 admission was not fully standardized, so this allowed for inclusion of all individuals with suspected COVID-19. Definitions and documentation within the EHR became more consistent as the study period progressed. As the burden of COVID-19 increased in NYC over the study period, most admissions were in fact for COVID-19.

## Conclusions

In this case series of patients tested for and hospitalized for COVID-19 at NYC’s public hospital system, male sex, older age, and certain chronic diseases were significantly associated with testing positive, hospitalization, and death from COVID-19, and racial/ethnic disparities were observed across all outcomes. These findings can help inform testing and mitigation strategies in other urban areas with similarly diverse populations.

## Data Availability

Data are from NYC Health + Hospitals' EHR system.

## Group Information

The New York City Health + Hospitals COVID-19 Population Health Data Team members include Spriha Gogia, PhD, MPH; Jonie Gordon; Richa Gupta, MPH; Nicole Hosseinipour, MPH; Laura Jacobson, MSPH; Remle Newton-Dame, MPH; Lauren Schreibstein, MA; Jesse Singer, DO; Jenny Smolen, MPH; Areeba Tariq, MS; Suharika Thotakura, MS; and Christine Zhang, MPA.

## Acknowledgments

We would like to acknowledge all of the patients and families in New York City who have been profoundly impacted by COVID-19 and the frontline clinical teams across New York City Health + Hospitals who have worked tirelessly in their care. We are particularly grateful to Dr. Mitch Katz and Dr. Machelle Allen for their clinical leadership during this extraordinarily challenging time. We also would like to thank Sharanjit Kaur for her assistance in manuscript preparation.

